# Single-cell RNA expression profiling of ACE2, the putative receptor of Wuhan 2019-nCoV, in the nasal tissue

**DOI:** 10.1101/2020.02.11.20022228

**Authors:** Chao Wu, Shufa Zheng, Yu Chen, Min Zheng

## Abstract

A novel coronavirus (2019-nCoV) was first identified in Wuhan, Hubei Province, and then spreads to the other Provinces of China. WHO decides to determine a Public Health Emergency of International Concern (PHEIC) of 2019-nCoV. 2019-nCov was reported to share the same receptor, Angiotensin-converting enzyme 2 (ACE2), with SARS-Cov. Here based on the public single-cell RNA-Seq datasets, we analyzed the ACE2 RNA expression profile in the tissues at different locations of the respiratory tract. The result indicates that the ACE2 expression appears in nasal epithelial cells. We found that the size of this population of ACE2-expressing nasal epithelial cells is comparable with the size of the population of ACE2-expression type II alveolar cells (AT2) in the Asian sample reported by Yu Zhao et al. We further detected 2019-nCoV by polymerase chain reaction (PCR) from the nasal-swab and throat-swab of seven suspected cases. We found that 2019-nCoV tends to have a higher concentration in the nasal-swab comparing to the throat-swab, which could attribute to the ACE2-expressing nasal epithelial cells. We hope this study could be informative for virus-prevention strategy development, especially the treatment of nasal mucus.

---

Severe infection by 2019-nCoV could result in acute respiratory distress syndrome (ARDS) and sepsis, causing death in approximately 2% of infected individuals^1^. Once contacted with the human airway, the spike proteins of 2019-nCoV can bind the surface receptors of sensitive cells, and mediate the virus to enter into target cells. Recently, Xu et.al. modeled the spike protein and indicated that Angiotensin-converting enzyme 2 (ACE2) could be the receptor for this 2019-nCoV^2^. Zhou et. al. showed that ACE2 is essential for 2019-nCov to enter HeLa cells^3^. These data indicated that ACE2 is likely to be the receptor for 2019-nCov. Once the putative receptor of 2019-nCoV is identified. An urging work is to identify the potential target cells of 2019-nCoV in the human body comprehensively.

Previous studies have investigated the RNA expression of ACE2 in 72 human tissues^4^. They found ACE2 is abundantly present in humans in the epithelia of the lung and small intestines. They also found ACE2 expression in the basal layer of the non-keratinizing squamous epithelium in nasal and oral mucosa and the nasopharynx. Yu Zhao et al. analyzed the single-cell RNA-Seq data of the lung tissues from eight donors and found that ACE2 mainly expressed in the type II alveolar cells (AT2)^5^. They found that ACE2 shows significantly high expression in AT2 of an Asian donor. However, the abundance of ACE2 expression in the upper respiratory tract needs further identification.

The upper airway epithelium, which is mainly composed of basal and apical cells (ciliated and glandular, and so on), ensures proper mucociliary function and can regenerate in response to assaults. Ordovas-Montanes J et al. proposed to use KRT5 as a marker of basal cells, and KRT8 as a marker of apical cells in the single-cell profiling of human respiratory epithelial cells^6^. Sandra Ruiz García et al. have profiled single-cell gene expression of airway epithelium from nasal brushing, bronchial biopsy, and turbinate^7^. Yu Zhao et al. showed the single-cell RNA-Seq data of the lung tissue from the Asian donor. In current work, we analyze ACE2 single-cell profiling in the tissue from different locations of the respiratory tract (nasal brushing, bronchial biopsy, turbinate, and lung tissue of the Asian donor).

We performed unsupervised graph-based clustering (Seurat version 3.1.4) on the single-cell RNA-Seq data of the nasal brushing, bronchial biopsy, turbinate, and the lung tissue of the Asian donor. We visualized the data using t-distributed stochastic neighbor embedding (tSNE) (Figure 1). Yu Zhao et al. use SFTPC as the marker of AT2. We didn’t detect SFTPC expression in the single-cell RNA-Seq data of nasal brushing, bronchial biopsy, turbinate. We found that KRT8 shows a wide expression in the cells of nasal brushing, bronchial biopsy, turbinate, and the lung tissue of the Asian donor. We used KRT8 as the marker of apical cells, and KRT5 as the marker of basal cells. We found KRT8 and SFTPC both mark AT2 in cells of the lung tissue of the Asian donor. We count the number of detected cells, cells with ACE2 expression, cells with KRT8 expression, and cells with both ACE2 expression and KRT8 expression in the single-cell RNA-Seq data of the nasal brushing, bronchial biopsy, turbinate and the lung tissue of the Asian donor (Table 1). We found that about 3.2% and 2.1% of KRT8-expressing cells in the nasal brushing and turbinate samples show ACE2 expression, which is comparable to the percent of KRT8-expressing cells in the Asian donor having ACE2 expression.

**Table 1:** The number of cells with ACE2/KRT8 expression in the tissues at different locations of the respiratory tract

| Samples | Total cells | ACE2-expressing cells | KRT8-expressing cells | ACE2-expressing and KRT8-expressing cells | The percent of KRT8-expressing cells with ACE2 expression |
| --- | --- | --- | --- | --- | --- |
| Lung/alveolus (The Asian donor) | 3855 | 95 | 1773 | 63 | 0.036 |
| Bronchial biopsy | 10795 | 24 | 3277 | 17 | 0.005 |
| Nasal brushing | 6645 | 163 | 5007 | 160 | 0.032 |
| Turbinate | 6438 | 106 | 4812 | 101 | 0.021 |

**Figure 1:**
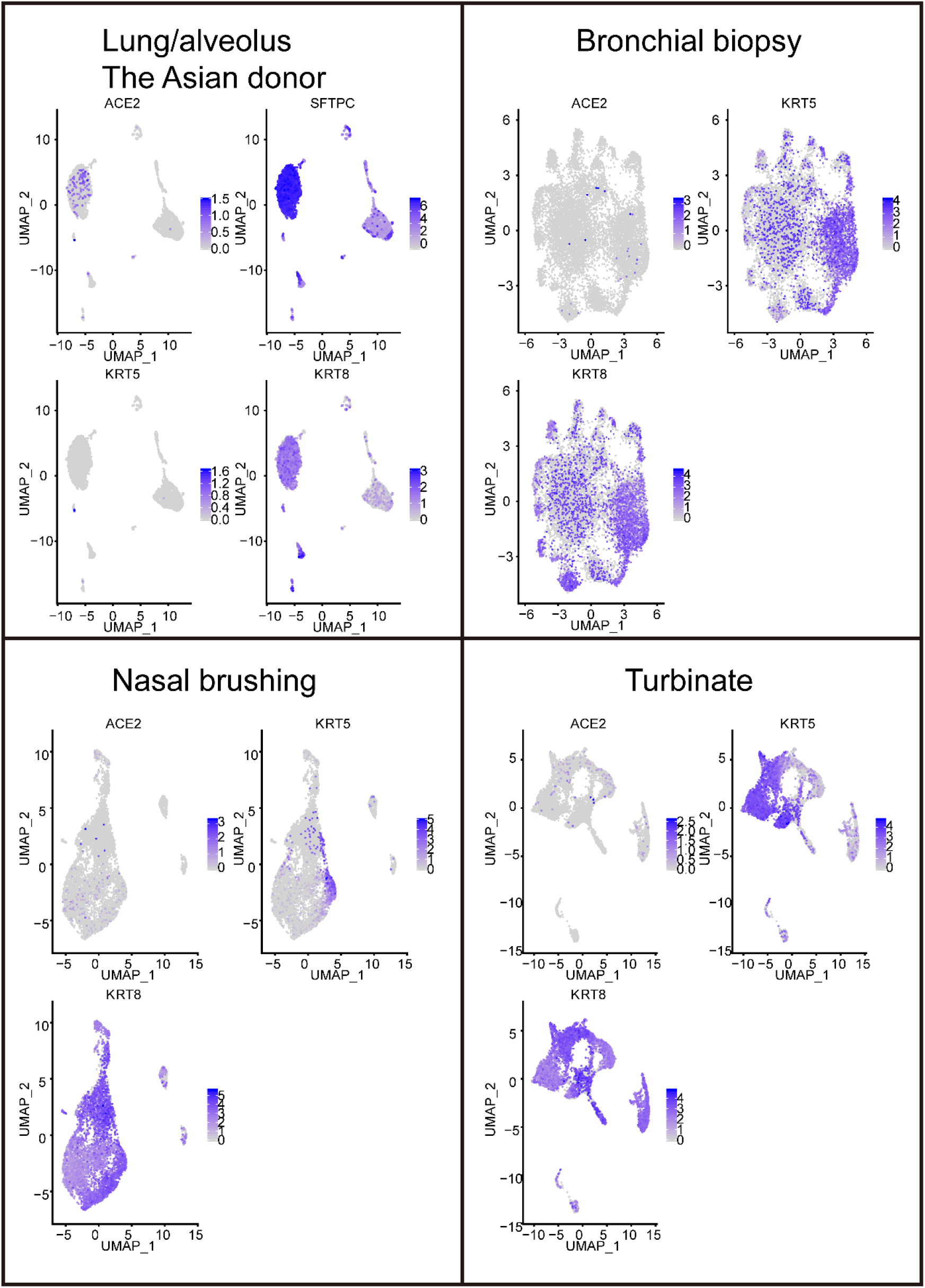
The tSNE of the cell population in the tissues at different locations of the respiratory tract The cells expressing ACE2, SFTPC, KRT5, and KRT8 are highlighted.

Seven were suspected as patients being infected 2019-nCoV in the First Affiliated Hospital, College of Medicine, Zhejiang University. The viral titers of 2019-nCoV in their nasal- and throat-swabs were detected by polymerase chain reaction (PCR) and measured by cycle threshold (CT). We found that 2019-nCoV is detected in his/her nasal-swabs, but not detected in his/her throat-swabs in two patients (patient W1 and Z1 in Table 2). In the patient Z2, 2019-nCoV is confirmed in his nasal-swab. But the PCR report of the throat-swab is suspected. In the patient Z4, 2019-nCoV is detected in both nasal- and throat-swabs. However, the CT value suggests that viral titers are higher in nasal-swab than in throat-swab.

**Table 2:** Detection of 2019-nCoV in the nasal- and throat swabs by PCR

| Patients | Throat-swab (CT value) | Nasal-swab (CT value) |
| --- | --- | --- |
| W1 | - | E:31.85; N:31.85; ORF:29.57 |
| W2 | - | - |
| W3 | - | - |
| Z1 | - | E:30.16;N:31.7;ORF:30.16 |
| Z2 | E:-;N:-;ORF:38.25 | E:34.41;N:39.02;ORF:35.17 |
| Z3 | - | - |
| Z4 | E:34.86;N:37.96;ORF:36.58 | E:31.51;N:33.22;ORF:32.36 |

-: No detection; N,E,ORFab1: N, E, and ORFab1 genes of 2019-nCoV.

Combining the results of ACE2-expressing cells in the nasal brushing and turbinate samples and the viral titers of 2019-nCoV in patients’ nasal- and throat-swabs, we infer that there exists a significant number of 2019-nCoV host cells in the nasal tissue. Decades ago, the basal layer of the nasal mucosa was reported to have ACE2 protein expression, which suggests that the candidate host cells of 2019-nCoV exist in nasal tissue. However, the analysis is at the bulk level and could not calculate the number of host cells. Our single-cell RNA-Seq analysis suggests that a significant number of candidate host cells exist in nasal tissue. China is suffering from the 2019-nCoV right now. Our results suggest that we should pay more attention to protect nasal tissue from 2019-nCoV infection.

## Method

The single-cell RNA-Seq data of lung/alveolus (the Asian donor) sample is downloaded from GSE122960; the single-cell RNA-Seq data of samples from bronchial biopsy, nasal brushing, and turbinate are downloaded from GSE121600. We filter out the cells 1-expressing less than 200 genes; or 2-highly expressing mitochondrial genes, in which mitochondrial genes’ reads account for more than 25% of the total reads. We filter out the genes expressing in less than 3 samples. Then, we conduct tSNE analysis on the cells with Seurat (3.1.4) in default mode.

Respiratory specimens (nasal- and throat-swabs) were collected to determine the amount of 2019-nCoV viral RNA by PCR analysis. According to the testing technology recommended by the National Health and Family Planning Commission of China, we detect the ORFab1, N, and E genes of 2019-nCoV. The detection limit of the ORFab1, N, and E RT-PCR assays was approximately 100 copies of RNA per mL.

The study was approved by Clinical Research Ethics Committee of the First Affiliated Hospital, College of Medicine, Zhejiang University (IIT20200012A).

## Data Availability

All the single-cell RNA-Seq datasets were downloaded from GEO database and the ID of the datasets were provided in the manuscript. Everybody can download them freely.

## Acknowledgements

This work was supported by the grants from the National Key Programs for Infectious Diseases of China (2017ZX10103008).

